# Modelling Semi-Random Human Mixing: A Modified Force of Infection in Population-level Epidemic Modelling

**DOI:** 10.1101/2025.08.14.25333622

**Authors:** Michael Lazarus Smah, Anna C. Seale, Kat S. Rock

**Affiliations:** Institute for Global Pandemic Planning, Warwick Medical School, Coventry, CV4 7AL, United Kingdom; System Biology and Infectious Disease Epidemiology Research, University of Warwick, Coventry, CV4 7AL, United Kingdom; Warwick Mathematics Institute, Coventry, CV4 7AL, United Kingdom; London School of Hygiene & Tropical Medicine

**Keywords:** Epidemic models, Random mixing, Clustered interactions, Basic reproduction number, Non-pharmaceutical interventions

## Abstract

Mathematical modelling can help inform public health policies; however, the assumptions about mixing impact results. Classical models typically assume everyone has an equal likelihood of interacting with everyone else, whereas in reality, people more frequently interact within smaller clusters, such as households, schools, or workplaces. In this study, we propose the *Semi-Random Mixing (SeRaMix)* approach based on network connectivity, where individuals have localised interactions with sparse external links. We derive an equation-based SeRaMix model (SeRaMix-EBM) with a modified force-of-infection function that preserves mathematical tractability while capturing more realistic mixing patterns. We derive its basic reproduction number (*ℛ*_0_) and, using numerical simulation, examine how cluster size and external connections shape outbreak dynamics compared to the classical approach. We validated the results of our SeRaMix-EBM against the stochastic, individual-based model (IBM) simulations. The results show that the SeRaMix-EBM is a good approximation for the IBM, but with much less computational complexity. Model simulations show that the predicted impact of interventions can be different between the classical and SeRaMix-EBM, depending on what type of intervention is considered. These findings indicate that semi-random mixing should be considered when creating models to inform policy, particularly for predicting outbreaks and the impact of non-pharmaceutical interventions.

## Background

Infectious disease outbreaks adversely affect society directly through impacts on health and, more broadly, through economic and social impacts. In this context, it is important to improve the understanding of the spread of pathogens among individuals to prevent or control outbreaks. Mechanisms to prevent severe disease or control the spread of infectious diseases include treatments, vaccination, and the use of non-pharmaceutical interventions (NPIs). NPIs aim to change social and behavioural patterns and reduce transmission by decreasing interactions and the risk of infection from these interactions. For emerging infectious diseases where pharmaceutical interventions are limited or not yet available, NPIs can reduce spread until other measures become available [1, 2, 3]. In either case, being able to predict the effectiveness of different interventions is important for outbreak response.

Mathematical modelling is an important tool used to advise policy-makers on the measures for prevention and control of infectious disease [4]. Analysis of mathematical models can give insight into the mechanism driving the spread of pathogens and help predict the course of the outbreak under different scenarios. Due to various factors, the robustness of compartmental disease models—where the population is partitioned into infection states such as susceptible and infected and the total number in each state is tracked—can be limited, for example, some models used during the COVID-19 pandemic were found to be more effective in the short-term prediction of the epidemic than in the long-term prediction [5]. Although some of the models were useful and had positive impacts on government policies during the pandemic, the validity and credibility of compartmental models have been debated [5, 6, 7].

A perpetual balancing act for modellers is to capture the complexity of epidemic processes while ensuring mathematical clarity and usability; this challenge in developing a theoretical mathematical framework that represents the outbreak scenario has been highlighted elsewhere [8]. This balancing would benefit from relaxing the assumption of random mixing in the compartmental models and overcoming the challenges of using detailed information on the network of contacts, for example by using methods such as the network structure [9, 10], variability in age [11], and geographical region (expert opinion in [12]). Notably, approximating network-based models into a simple mathematically tractable compartmental modelling framework had been suggested and attempted in the literature [13]; this approach considered the non-random contacts per individual, as each person makes contact with the same number of neighbours per unit time. The main network properties include the average number of contacts per individual, the variance in the number of contacts, and the clustering (the ratio of the number of connections in a set of three nodes with edges between every pair to the total number of three nodes with at least two edges among them) of the network. The clustering structure of networks was found to be one of the key parameters determining the dynamics of epidemics; however, the difficulty in measuring the clustering was acknowledged, hence the need for research in this area.

Understanding the formulation of the transmission term could help efforts towards improving our understanding of epidemic dynamics. The transmission term is a key part of epidemic modelling, and the right description and formulation of it play a role in better predicting and estimating the quantitative impact of different possible interventions. Although, several formulations for the transmission term have been proposed and discussed in the literature [14, 15, 16], the prominent among them for human-to-human transmission is the classical frequency-dependent transmission model [17, 18, 19] which assumes each person has the same average number of daily contacts regardless of the population size.

There are two main constituents of the classical “susceptible-infected-recovered” (SIR) model transmission term; firstly, the effective contact rate, *β* which is a product of the rate of contacts per unit time, *c*, and the probability that the contact with an infected person leads to the transmission of the pathogen, *µ*, and the probability that a contact will be made with an infected person,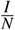, where *I* is the number of infected individuals, and *N* is the total number of individuals in the population [14, 20]. Thus, the rate of creating new infections in the population is 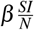, where *S* the number of susceptible individuals *S*.

A key assumption in the classical model is that of random mixing that everyone is equally likely to meet everyone else [7], however, in practice, there are many nuances in the contact patterns of individuals in a population, some of which are typically addressed by expanding the classical model to incorporate heterogeneous mixing patterns such as by using a “who-acquires-infection-from-whom” (WAIFW) matrix linked to data on age-depending mixing [21, 22]. Yet even within demographic or other groupings, these model formulations generally still retain the ability for anybody to mix with anyone else on any day, albeit with weighted probability. Others have used the approach of developing individual-based models (IBMs), which can explicitly use probabilities and networks to capture this type of mixing behaviour, but IBMs can be computationally slow and analytically intractable [23, 24].

It is established knowledge that non-random mixing influences the spread of pathogens, and some experts have suggested that incorporating social networks into epidemic models would be better than the assumption of random mixing [9, 12]. A major challenge to achieve this is the complexity of obtaining detailed information on the web of the network of contacts [10] and obtaining a population-level model that better accommodates this.

This study aims to develop a mathematically tractable equation-based model (EBM) that approximates the effect of the semi-random mixing inherent in human contact networks—where each individual draws daily contacts from a fixed pool of potentially recurrent intra/inter-cluster connections—by modifying the classical force of infection term. Specifically, we will:

i. Modify the classical frequency-dependent force-of-infection to create our novel semi-random mixing (SeRaMix) EBM, which blends within-cluster and external contacts in a single expression.
ii. Ensure analytical tractability by choosing a functional form that permits derivation of key epidemic quantities (e.g., basic reproduction number, *ℛ*_0_).
iii. Analyse how cluster size and external connectivity shape transmission dynamics—comparing against the standard classical homogeneous mixing model.
iv. Validate the new SeRaMix-EBM approach through analogous IBM simulations of an SIR-type infection, demonstrating how semi-random mixing alters outbreak timing, peak size, final size, and the analysis and model recommendations of NPIs.

Our goal is to create a model that is both more realistic than the classical EBM (with SeRaMix capturing how people interact non-randomly) yet mathematically simple enough to analyse in a reproducible and rapid manner, unlike an IBM. This approach could improve infection modelling and help better plan outbreak responses.

## Methods

We consider a population of *N* individuals, distributed across *K* clusters. Each individual in cluster *k* of size *n*_*k*_ is connected (not to be confused with making contact) to the other *n*_*k*_ −1 members of their cluster. In addition, each person has *x*_*k*_ external connections randomly chosen from the other *K*− 1 clusters. For each person, (*n*_*k*_−1) + *x*_*k*_ is the number of their connections from which daily contacts are randomly selected. We want to measure how saturated the population is in terms of connectivity of people (accounting for how likely everyone will make contact with everyone else). We consider the average cluster size to be *n* and the average external connection to be *x*.

### Human Potentially Recurrent Contact Networks

Human interaction networks are neither random nor uniform [25, 26, 27]. Instead, they are naturally shaped by shared locations of interaction and social structures, such as families in households, students in classrooms, employees in office spaces, religious gatherings, sports teams, dormitories, etc. These community-based clusters represent subgroups in the larger human contact network, where individuals are more likely to repeatedly interact with members of their immediate group (referred to here as a cluster). However, individuals are not strictly isolated within these clusters; for example, a student might be friends with someone from another class, a worker might socialise with someone in a different office, and neighbours form friendships across households. Thus, each person possesses a set of potentially recurrent contacts—individuals from both within their cluster and a set of external connections out of which daily contacts may be randomly made. These are not random encounters across the entire population, but structured selections based on familiarity, proximity, or habitual contact.

As interactions are chosen randomly, but from a fixed pool of potentially recurrent contacts, rather than the entire population, the potentially recurrent contact network (PRCN) as depicted in Fig. 1 emerges, characterized by: a high likelihood of daily contacts within the same cluster (household, class, etc.), and the number of external connections. In this approach, we assume that a person’s contacts for the day are not entirely random and can only be selected from a defined pool of recurrent contacts; however, there is randomness between days about which subset of the recurrent pool of people is contacted. Consequently, the spread of infectious diseases may be influenced by the size of potentially recurrent contacts per individual.

**Figure 1.**
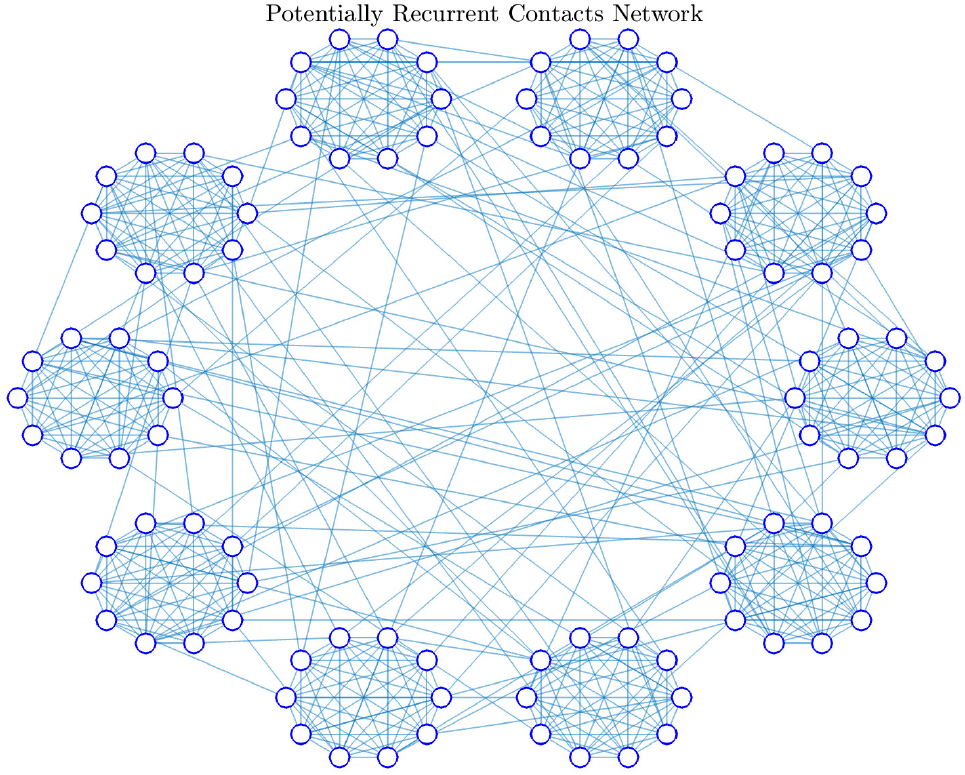
Network visualisation of 100 individuals organised in 10 equally-sized clusters, with intra-cluster connections and inter-cluster (external connection) links of 1 per person, illustrating the PRCN structure. Each white circle represents an individual, and the solid lines represent connections between pairs of individuals.

Due to the limited connections with the entire population, once a pathogen enters a cluster, it is likely to circulate locally within a cluster before escaping. As such, some clusters may be affected early, while others remain untouched for longer periods or throughout the outbreak. Populations with the same size but different clustering structures (e.g., two populations where one has a high average household size vs. another with smaller average household sizes) may experience different dynamics of spread—including in duration, peak height of infection, and the epidemic final size. The variables and parameters notation used in this study and their descriptions are given in Table 1.

**Table 1.**
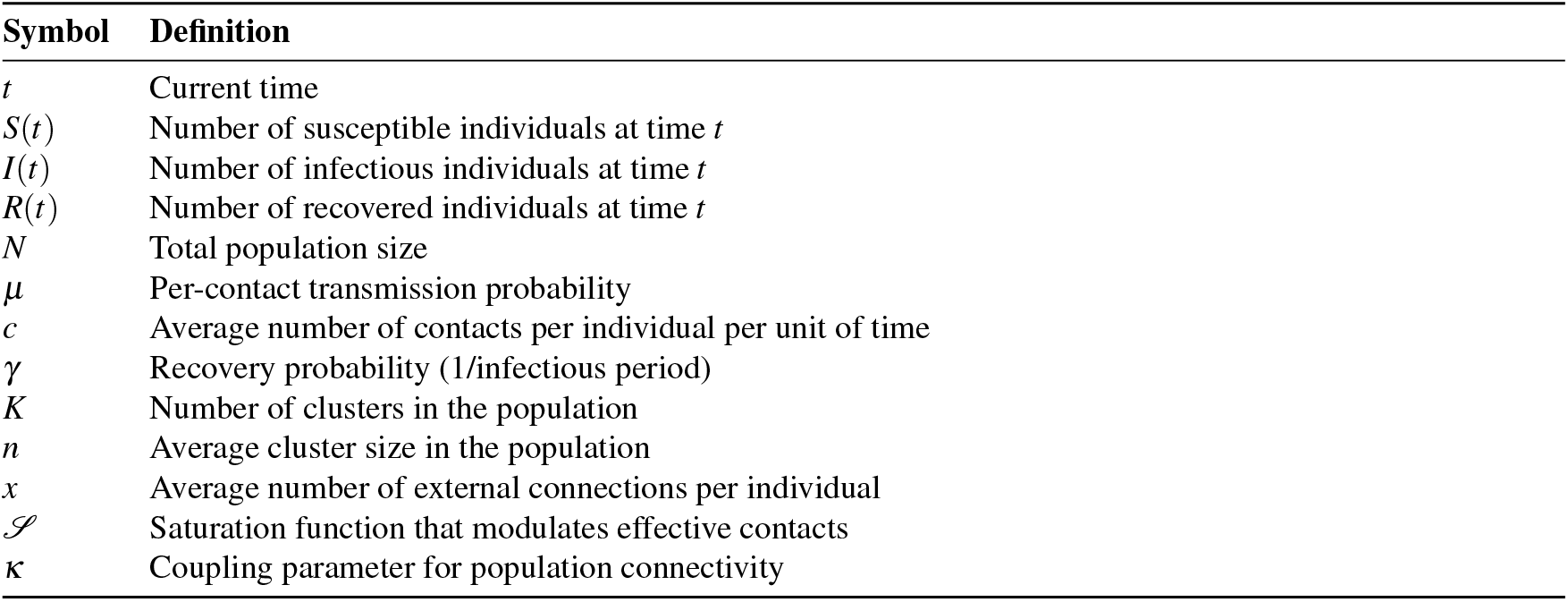
Model variables, parameters, and definitions

To measure the *saturation* of the connections, we use a probabilistic model based on percolation theory [28, 29] and network connectivity [30, 31, 32] (see the Supplementary Information for the full model formulation). We utilise two known characteristics of Erdős–Rényi (ER) random graphs: the giant component and the full connectivity threshold, which leads to our EBM approximation:

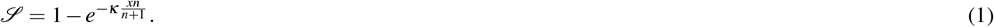

which we refer to as the saturation function. In this expression, *κ* is a coupling constant for population connectivity, used as a correction to overestimation that may arise as a result of a simplifying assumption in formulating the saturation function. When *κxn*≪*n* + 1, *𝒮* ≈ 0, it corresponds to fragmented clusters with little or no interactions between clusters. When *κxn*≫*n* + 1, *𝒮* ≈ 1, matching the full connectivity regime. The transition between these extremes mirrors real-world connectivity that we aim to quantify as suitable for different average cluster sizes, *n*, and the average per-capita external connections, *x*. This ratio quantifies how far the population is from achieving contact saturation.

### Modelling disease transmission

The derivation of the saturation function is intended to utilise known principles to obtain an equation-based, population-level quantification of the impact of semi-random mixing in the population, rather than engaging in a full (numerical) analysis of a contact network (such as Fig. 1).

The saturation function formulated here captures the essential components of connectivity-driven processes, making it suitable for modelling and analysing phenomena such as epidemic spread, where the efficiency of transmission depends heavily on the underlying connectivity structure. In general, we assume that the likelihood that any random person will transmit infection to any other random person in the population is weighted by the saturation function. This is intended to give a population-level approximation of the limiting consequence of sustaining the pathogen within a limited social circle for some time before transmitting to other individuals in other clusters directly or through a chain of connections. The infection in the population of *N* individuals, distributed in *K* clusters of average size *n*, where each person has an average of *x* external connections randomly chosen from the other *K*−1 clusters is a product of (i) the average number of contacts per time per person, *c*, (ii) the probability that contact with an infected person will lead to transmission, *µ*, (iii) the prevalence of disease in the population at 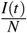, and (iv) the cluster saturation index, 𝒮. That is, if the proportion of infected people in the population is 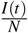, then the overall likelihood that a person will meet an infected person in the population depends on how likely everyone will meet everyone else, 𝒮.

The first three quantities in the force of infection described above are well known in the field of epidemic modelling (see [14, 15, 16, 17, 18, 19]). The proposed modification in this work is the inclusion of the saturation function so that the probability that a random contact made (which is likely from the same set of people for a long time) will be with an infected person is a function of the disease prevalence and saturation function, 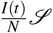.

To write down the equation of disease evolution in the population of *N* individuals, we consider that the infected individuals recover at rate *γ*, to write the system of ordinary differential equations (ODEs) for the SIR-type infection as:

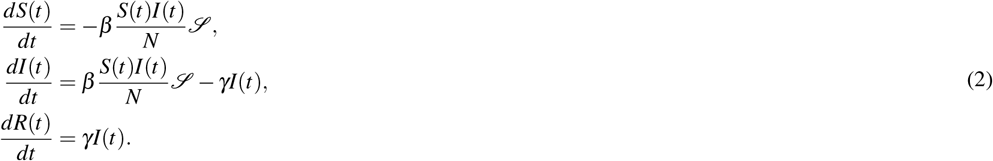

In this model, the force of infection on susceptible people is given by:

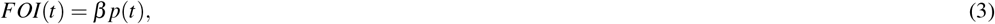

where 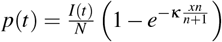 is the per-contact probability of choosing an infected person.

### Special (extreme) cases

i. If *x* = 0, implying that there are no external connections, we have *FOI*(*t*) = 0. Although members of the same cluster are expected to be exposed to or infected by the infected member of their cluster, the infection will die out within the cluster; as such, the population-level likelihood of meeting the infected is drastically limited.
ii. Similarly, if *n* = 1, implying that there is only a single person in each cluster, and for small *x* (limited external connections), we have *FOI*(*t*) ≈0. This is because if there is no other person in the cluster to interact with, and there are limited connections with other clusters, then the per-contact probability of meeting an infected person is small.
iii. If the number of individuals in the cluster approaches the population size (*n→N*) and/or the number of external connections is close to the population size (*x→N*), then *FOI*(*t*) 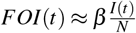. In this result, the per-contact probability of meeting an infected person corresponds to the term used in the classical frequency-dependent transmission models.

We now reformulated the force of infection (details in the Supplementary Information) in terms of *µ* (which is pathogen-dependent), the contact rate *c*, and the per-contact probability of choosing an infected person, *p*(*t*). This approach would enable us to distinguish the behavioural and social constituents (*c, n*, and *x*) in the force of infection from the biologically innate quantity, *µ*, in the analysis of non-pharmaceutical intervention. Based on this, the probability that a susceptible person will be infected after contact with *c* persons of unknown infectious statuses at any time *t* (the force of infection) is given as:

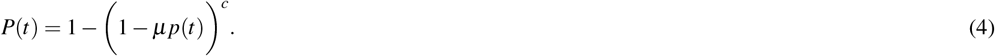

The discrete-time SeRaMix equations for this SIR-type infection are thus written as:

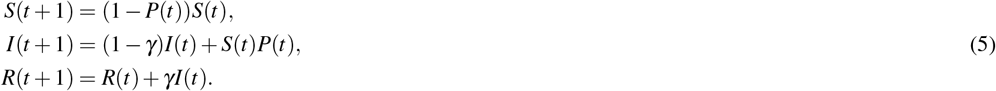

We additionally derived the basic reproduction number (detailed derivation in the Supplementary Information) to obtain the analytical expression representing the average number of secondary infections from a typical infected person in a totally susceptible population.

### Model validation

To validate this formulation and estimate *κ* in our SeRaMix-EBM, we used an IBM to simulate the spread of an epidemic in clustered populations (detailed approach and algorithms are in the Supplementary Information). We use an IBM because it provides the closest modelling representation of real-world interactions, where each individual possesses their unique cluster neighbours and external connections, and as such, is a good method to validate our EBM approximation. Additionally, unlike our EBM approximation, it does not require the value of *κ* to simulate the outbreak. Since the aim is to observe the effect of semi-random mixing for different population structures, other individual-based heterogeneous characteristics are not considered here, except for connectivity. We will compare the infection results of our SeRaMix-EBM with the results of IBM. Since we are not using any real data in this work, we will consider the outbreak results from IBM to represent “real” data to fit the coupling parameter *κ* and compare the disease dynamics for different *n* and *x*.

For IBM, we generated *N* = 1, 000 individuals and randomly assigned each to a cluster to mimic random local interactions at work or home. For each individual, we assign *x* randomly selected individuals from other clusters. For each individual, we assigned their *n*−1 cluster neighbours and the *x* randomly selected individuals as their social circle (these are the potentially recurrent contacts of each individual) so that each person has *n*−1 + *x* connections out of which they randomly select their *c* daily contacts at each time step. This implementation is presented in **Algorithm Box 1** of the Supplementary Information.

Firstly, we simulated the IBM with varying network structures with average number of external connections, *x*, and average cluster sizes, *n*. We used the infection results obtained from IBM as synthetic data to estimate the value of *κ* in the discrete-time EBM. To do this, we employed non-linear optimisation techniques using MATLAB and following the steps in **Algorithm Boxes 2 and 3** of the Supplementary Information. We used the incidence (daily new infections) data to fit the EBM. This choice of incidence data in model fitting appears in the literature (see, for example, [33, 34]), and has advantages over other metrics, e.g. incidence is more commonly reported than prevalence, and cumulative incidence has a lack of independence between time points that may present challenges in parameter estimation as each point includes all previous cases. Both the EBM and IBM simulate infection spread over a structured population with fixed parameters: total population size *N* = 1000, per-contact transmission probability *µ* = 0.18, average contacts per individual *c* = 4, and daily recovery probability *γ* = 0.16. We chose the average cluster size *n*, and the number of external connections *x* to run the IBM model, and stored the number of new infections computed at each time step. We used six values for the external connections, (*x* = 1, 2, .., 6), and four values for the cluster sizes, (*n* = 1, 2, .., 4). For each *n, x* combination, we ran 1,000 realisations, resulting in 1,000 datasets. Thus, there are 24,000 distinct synthetic datasets used to fit the model. The reason for the choice of 24,000 datasets obtained from the different *n* and *x* is to estimate a *κ* value that is more or less a representation of a wide range of parameters and possible outbreak trajectories. Given the synthetic time series data, we defined an objective function as the sum of squared errors (SSE) between the EBM predictions and synthetic IBM data.

For each data set and time *t*, the residual (detailed algorithm for computing this is in **Algorithm Box 3** of the Supplementary Information) is:

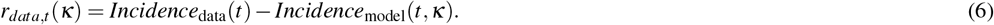

The objective function is the sum of squared residuals across all datasets and time points, given as:

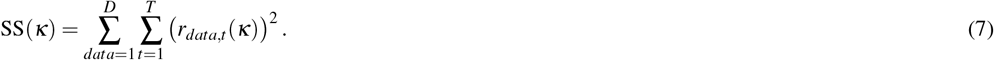

where *D* is the number of datasets and *T* is the number of time points.

This function was minimised using optimisation algorithms outlined in **Algorithm Box 2**. We used the `lsqnonlin` from MATLAB’s Optimisation Toolbox to estimate the coupling parameter *κ* in the discrete-time SIR-type model. The `lsqnonlin` is designed to minimise the norm of a vector of residuals and is particularly suitable for least-squares problems [35]. We defined a residual function (equation (6)) that simulates the SIR dynamics and computes the element-wise differences between the EBM-predicted and the IBM incidence data over time. These residuals were concatenated into a single vector, which was passed to `lsqnonlin` along with an initial guess for *κ*, and non-negativity constraints.

Finally, to assess how the SeRaMix-EBM differs from the classical EBM (without semi-random mixing) we performed additional simulations. In both models, we set *I*(0) = 1, with all other people being susceptible initially. To demonstrate that the classical EBM can be made to produce the same or similar results as our SeRaMix-EBM, we modified the average number of contacts *c* in addition to setting *x* = *n* = 1, 000 so that *c*≈2.7. We simulated two different models; the classical EBM used the baseline parameter values and *c*≈2.7 and *x* = *n* = 1, 000, and our SeRaMix-EBM used *c* = 4, *x* = 2, the fitted *κ*, found by fitting to the IBM, and *n* = 5. Next, we introduced a 50% reduction in the number of contacts for both the classical EBM and the SeRaMix-EBM on day 15 of the simulation and observed a change in the infection dynamics.

## Results

The analytical expression of the basic reproduction number, ℛ_0_ derived using this model is given as:

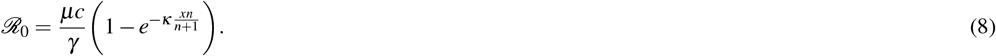

Similar to other epidemic models, the ℛ_0_ in this model depends primarily on two epidemiological quantities: the infectious probability *µ* and the recovery probability *γ*. However, this model reveals the additional dependence of ℛ_0_ on the saturation function, which is a function of the average cluster size in the population *n*, the average external connections *x*, and the coupling constant *κ*.

We computed the numerical value of the ℛ_0_ in Equation (8) to visualise the effect of different average cluster sizes and external connections per person on the ℛ_0_, given that everyone has the same average number of contacts *c*, the same disease with the same biological characteristics in terms of *µ, γ* is introduced (Figure 2).

**Figure 2.**
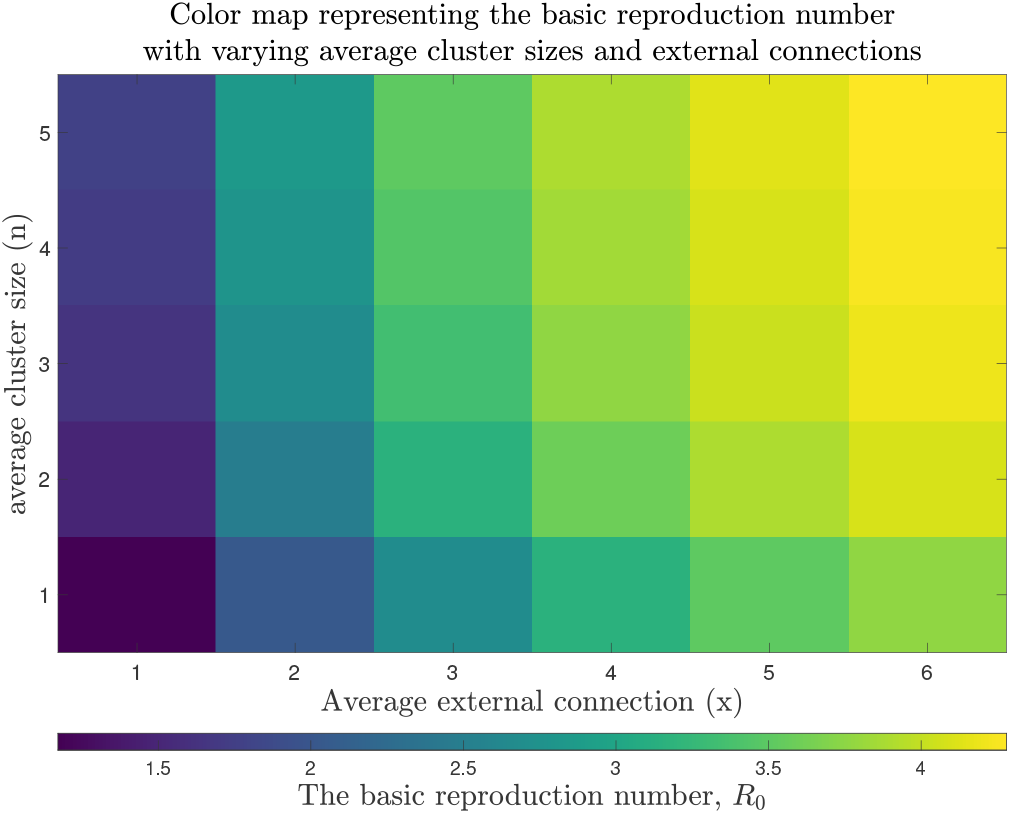
Heatmap of the basic reproduction number (ℛ_0_) as a function of average cluster size (*n*) and external connections (*x*). The color intensity represents the value of ℛ_0_ computed using the semi-random mixing model, with cluster sizes ranging from *n* = 1 to *n* = 5 and external connections from *x* = 1 to *x* = 6. All other parameters are: transmission probability *µ* = 0.18, recovery rate *γ* = 0.16, average contacts *c* = 4, average cluster size *n* = 5, and *κ* = 0.6031. The results demonstrate how both smaller cluster sizes (*n*) and fewer external connections (*x*) reduce ℛ_0_, highlighting the combined effect of local clustering and inter-cluster restrictions on epidemic potential. The colorbar indicates ℛ_0_ values, with darker colors representing lower transmission potential.

For the model fitted to synthetic data, the estimated value of the coupling constant is *κ* = 0.6031. This fitted value of *κ* was used to perform simulations of the model and compare the results with the synthetic data used to fit the model and plotted in Figures 3 and 4. The aim is to see how well the model would produce the data for a range of average cluster sizes and external connectivity. The model demonstrates consistent performance across most parameter combinations, with particularly good agreement at intermediate *x* values, but less good agreement for very low external connectivity (*x* = 1). The shared *κ* estimation approach ensures parameter consistency while accommodating connectivity variations.

**Figure 3.**
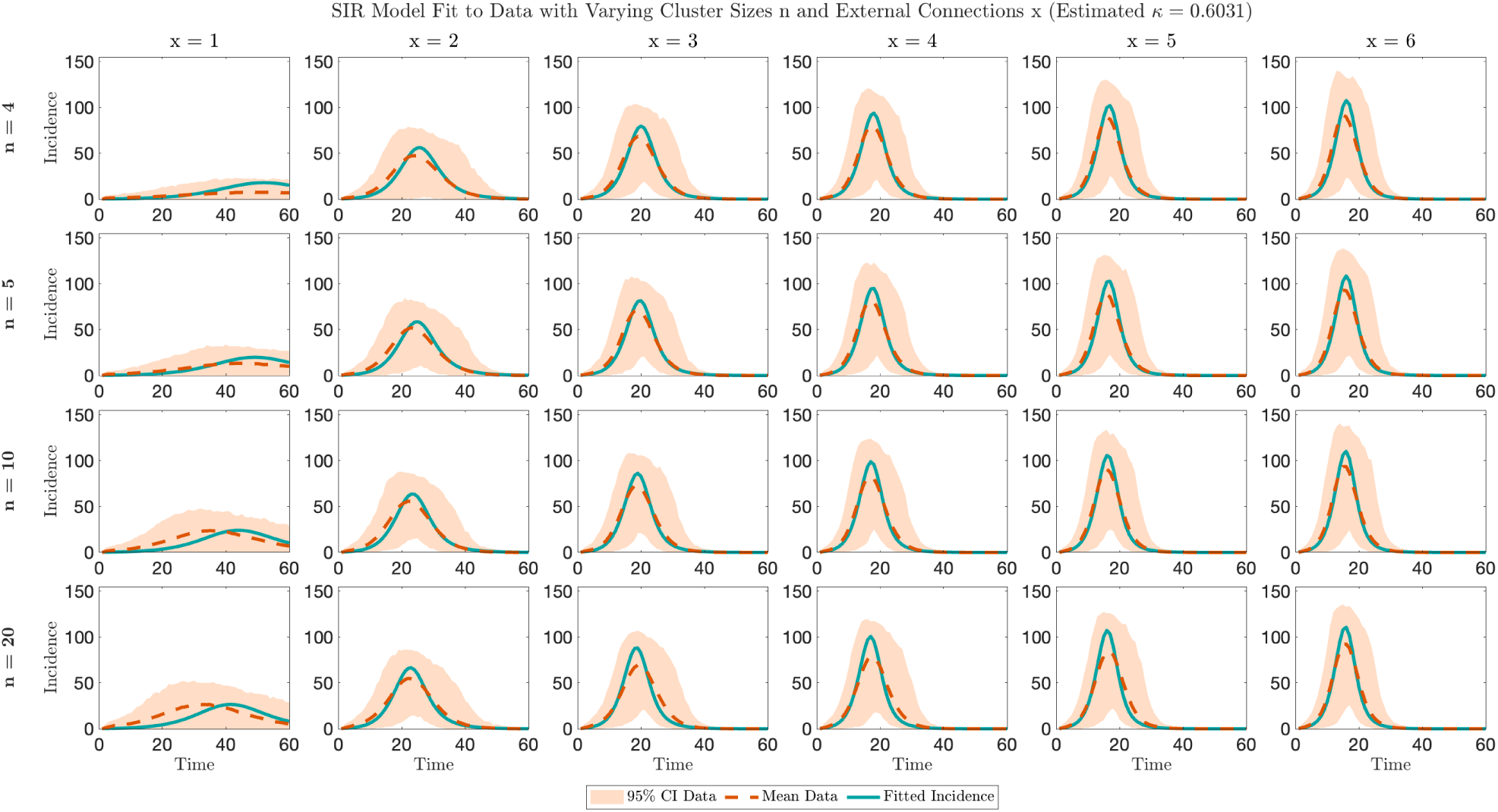
Comparison of EBM fits to the synthetic incidence data from the IBM realisations across varying cluster sizes (*n*) and per-capita external connectivity parameters (*x*). The estimated parameter *κ* was simultaneously fitted to all synthetic datasets. Each subplot shows the synthetic data (shaded region-95% C.I) and EBM fit (solid line) for: (a) cluster sizes *n* ∈ {4, 5, 10, 20} (rows), and (b) connectivity parameters *x* ∈ {1,…, 6} (columns).

**Figure 4.**
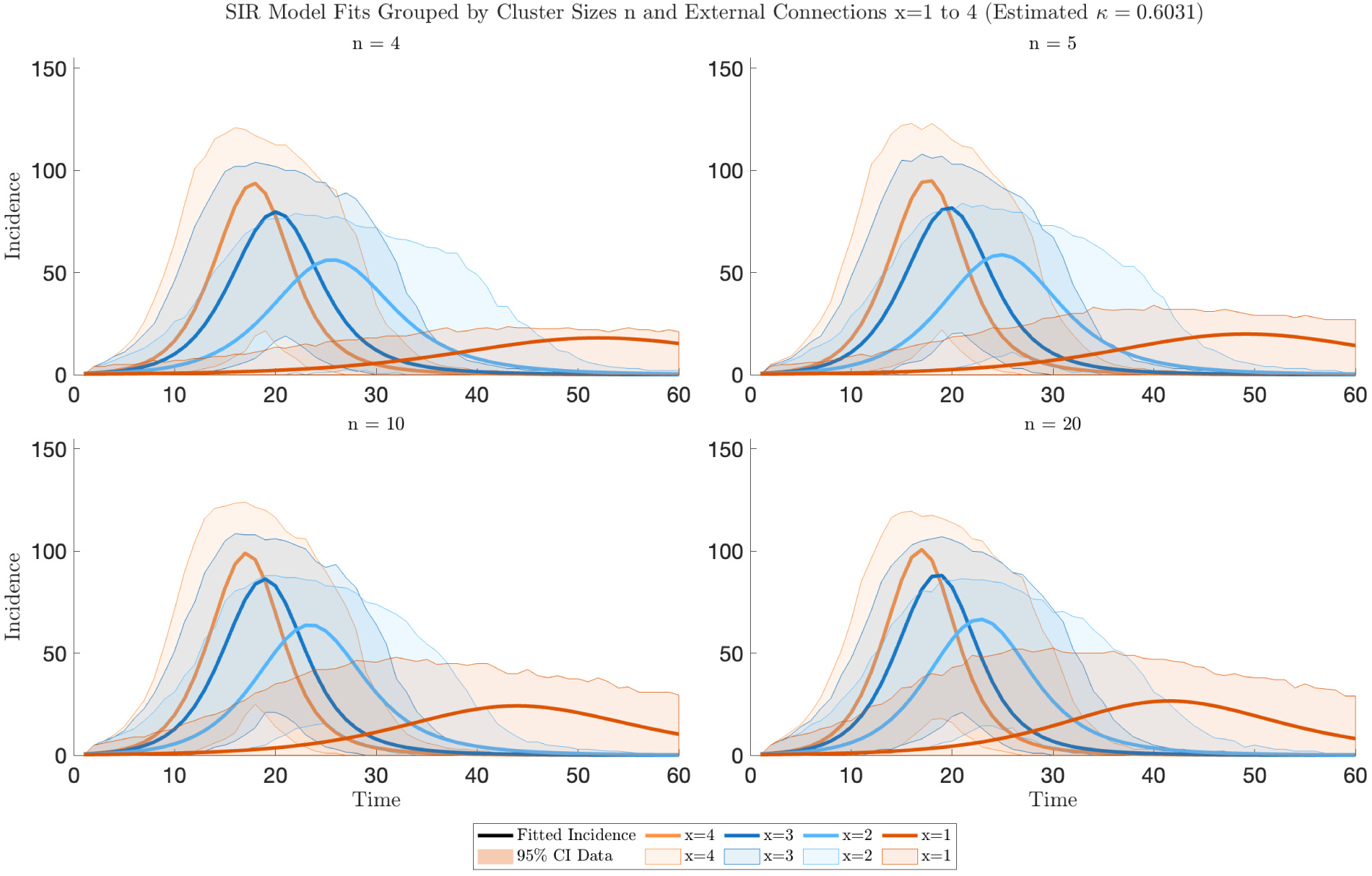
SeRaMix-EBM compared to the synthetic IBM data grouped by cluster size *n* using the globally estimated *κ* (all EBMs use the same value). Each subplot shows infection dynamics for all per-capita external connectivity (*x* = 1, …, 4) at: (a) *n* = 4, (b) *n* = 5, (c) *n* = 10, and (d) *n* = 20. For each *n* and *x*, 1,000 datasets have been used. The synthetic IBM data are represented by the shaded 95% prediction interval and dashed mean line. The EBM simulations are shown as the solid lines.

Figures 3– 4 in general show that the our SeRaMix-EBM can fairly well produces the dynamics obtained using the IBM approach with semi random mixing, and show that the smaller the average cluster size, the lower the epidemic peak height and the longer the duration of the outbreak, and vice versa. However, the peak infections for the EBM simulation appear higher than the mean of the synthetic data generated from IBM simulations. This result is not surprising, as the phenomenon where a deterministic model is not strictly the mean of its stochastic counterparts appears in the literature [36, 37, 38, 39]. The agreement between the EBM developed in this work and the corresponding IBM shows the validity of the model, particularly in modelling the semi-random mixing effect.

The validation of the SeRaMix-EBM approximation allows us to use the deterministic version for analysing other behaviours of the model, and counterfactual analysis of some NPIs. We performed numerical simulation of the SeRaMix-EBM (5) to show the effect of different average cluster sizes on the spread of infectious diseases (Fig. 5). We set the average cluster size *n* = 1, 000 corresponding to the number of clusters *K* = 1, implying that everyone interacts within the same cluster, and observed that the result corresponds to the classical EBM. However, if the cluster size is less than the population size, accounting for a more realistic scenario that people do not always interact within one cluster, the higher the cluster size, the higher the epidemic peak and the shorter the duration of outbreaks (Fig. 5 even if the same pathogen is introduced in different populations with the same average contacts per unit time per person.

**Figure 5.**
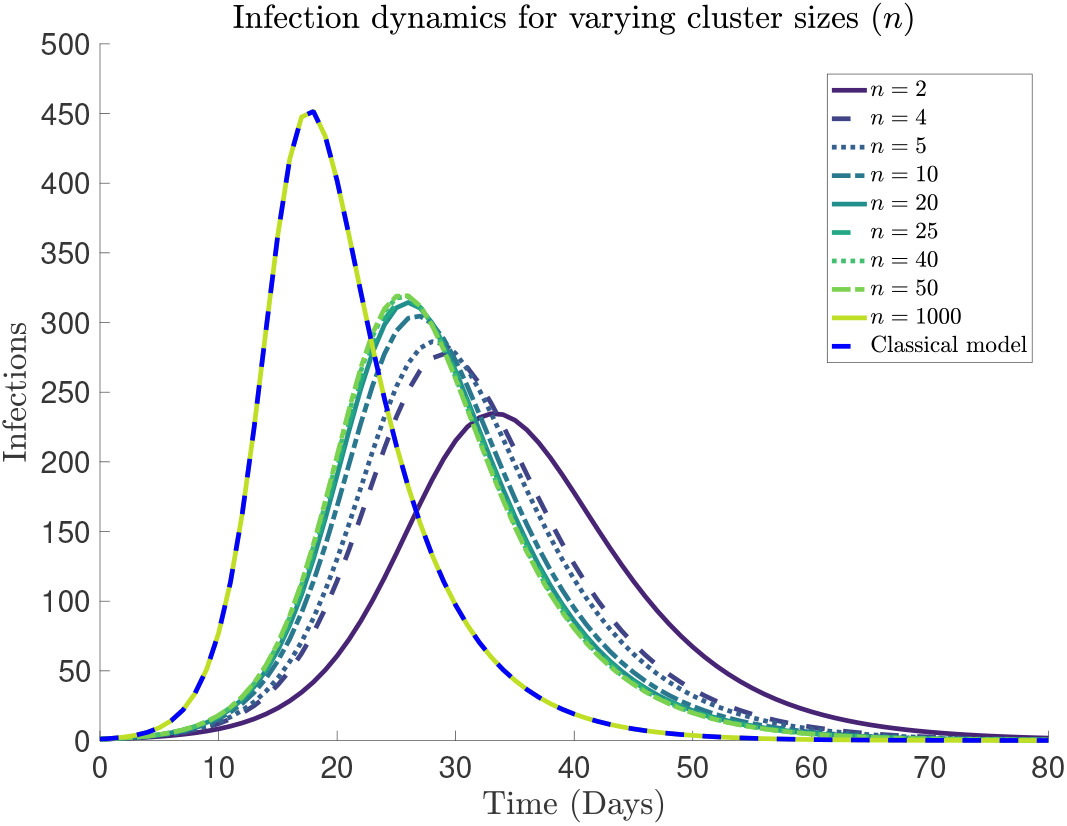
Infection dynamics for varying cluster sizes (*n*) compared to the classical epidemic model. The figure shows the evolution of infected individuals (*I*) for different cluster sizes ranging from *n* = 2 to *n* = *N* = 1000, where *N* represents the total population size. All other parameters are: transmission probability *µ* = 0.18, recovery rate *γ* = 0.16, average contacts *c* = 4, and *κ* = 0.6031. The classical model (dashed blue line) assumes homogeneous mixing (*n* = *N*). All simulations begin with a single infected individual (*I*(0) = 1) in a susceptible population (*S*(0) = 999). The results demonstrate how smaller cluster sizes lead to slower epidemic spread due to the semi-random mixing effect, where transmission is constrained by limited contact opportunities within and between clusters.

The comparison of the classical EBM and SeRaMix-EBM (although with different baseline parameter values in terms of *c, x*, and *n*) resulted in the same value of ℛ_0_ = 2.85 and the same infection dynamics (Fig. 6 without contact reduction). Unlike the classical EBM, this new SeRaMix-EBM allows us to explicitly target *x* as an NPI that corresponds to reducing visitation between members of different clusters, and *n*, which could mean reducing cluster sizes. If clusters are seen as households, reducing household sizes may not be feasible; however, if clusters are defined as in workplaces or schools, then reducing the cluster sizes would mean reducing the number of individuals in offices, classrooms, etc. These two parameters would aid counter-factual analysis of NPIs that are not readily available using the classical EBM (see Fig. 6).

**Figure 6.**
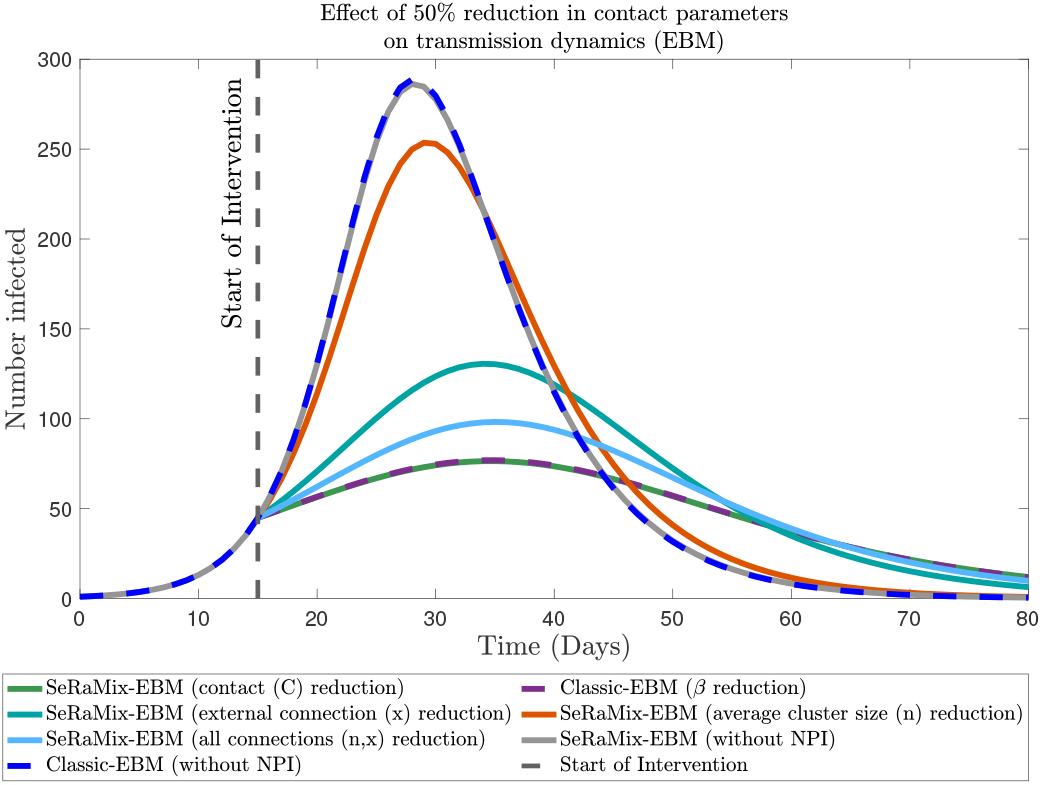
Comparison of different non-pharmaceutical intervention (NPI) strategies in both the semi-random mixing equation-based model (SeRaMix-EBM) and classical equation-based model (Classic-EBM). Both models were calibrated to yield identical baseline transmission potential (ℛ_0_) and epidemic size by adjusting contact parameters (SeRaMix-EBM: *c* = 4, *x* = 2, *n* = 5, *κ* = 0.6031; Classic-EBM: *c*≈2.7, *x* = *n* = 1000). The figure shows the number of infected individuals over time for SeRaMix-EBM and classic-EBM with NPIs, and both models without NPIs (grey and blue dashed, respectively). The black dashed vertical line indicates the start day of the intervention. All intervention types represent a 50% reduction in their respective contact parameters, corresponding to a reduction in the number of daily contacts, *c*, external connections, *x*, cluster size, *n*, or all connections (*n, x*).

## Discussion

This study developed a modified compartmental model that accounts for semi-random mixing in structured populations by introducing a transmission term weighted by a ‘contact saturation function’ which depends on the cluster size (*n*) and external connections (*x*). The model bridges the gap between fully homogeneous mixing (analytically tractable classical models) and computationally-expensive non-random mixing approaches, capturing the interplay of potentially recurrent contact networks observed within households and places of work/school and global connectivity.

The modified transmission term approximates the dynamics from IBM simulations well for a range of mixing structures (Figures 3–4), where contacts were randomly drawn from local clusters and external connections. The heatmap of ℛ_0_ (Figure 2) showed that both smaller clusters (*n*) and fewer external connections (*x*) reduce transmission potential. When *n* = 5 and *x* = 2 (chosen as our baseline parameterisation for SeRaMix-EBM), ℛ_0_ matched the classic model’s ℛ_0_ when the latter assumed a lower contact rate (*c* ≈ 2.7). This implies that *semi-random mixing inherently suppresses epidemic spread* as expected in reality, especially with moderate external connectivity.

The transmission term in different epidemic models takes different forms. Even in the most accepted equation-based transmission models, there are underlying assumptions to reduce the complexity of mathematical analysis. The advancement in computational techniques has made it possible to relax some assumptions and account for some of the complexities associated with human interaction (in particular those that would normally only be captured by approaches such as IBMs). However, using an EBM approach like the one presented in this study would be useful to give analytical insights into the parameters governing the behaviour of epidemic models and allow for rapid simulation.

The classic EBM produced the same epidemic dynamics as our SeRaMix-EBM in the absence of NPIs, by modifying the contact rates; however, the novel SeRaMix-EBM presented in this study enables the evaluation of realistic NPIs that target specific network properties (Figure 6), unlike the homogeneous contact reduction of the classical EBM. The semi-random mixing framework can model cluster-specific measures (e.g., workplace/school control through *n* reduction) and visit restrictions (via *x* reduction) that align better with different real-world interventions, rather than assuming uniform contact reduction across all individuals.

While classical models generalise outbreak trajectories for specific pathogens, our work emphasises how *network structure modulates epidemic shape* even when average contacts and biological characteristics of the pathogens are held constant. Smaller clusters slow early exponential growth (Figure 5), buying time for interventions.. Figure 5 shows that the smaller the cluster size, the slower the spread and the lower the peak of infections, and vice versa. This supports the strategy employed in England during the COVID-19 pandemic [40] where household members form a bubble of no more than two households where interactions are restricted to the members of the bubble. Hence, asking people to continuously meet with the same (smaller) set of individuals at home and at work would reduce the peak height of infections (Figure 5), and could essentially reduce the overwhelming of the healthcare system.

The ideas of modelling the spread of infection where a population is subdivided into smaller populations with the populations connected through the mobility of people (metapopulations) are available in the literature [41, 42, 43]. However, unlike the metapopulation that requires epidemiological information on the distinct sub-populations, this study is aimed at sub-populations like a town with many households, a school with different classes, a building with different office spaces, etc. where specific information on the populations infected or susceptible for example are not necessarily available at the local cluster level but where this information available at the population level can be harnessed to analyse the effect of the multi-cluster connectivity and non-random mixing in pathogen transmission.

This study demonstrates that classical EBM is a special case when *K* = 1 (single cluster population), and that the proposed SeRaMix framework is a generalisation of existing approaches. This unification provides new insights into when traditional models may over-or under-estimate transmission risks in structured populations. The framework provides a principled way to assess cluster-targeted strategies (e.g., household isolation, workplace closures) that are increasingly relevant in modern infection control.

The analytical derivation of ℛ_0_ reveals explicit dependence on both biological parameters (*µ, γ*) and network structure (*n, x, κ*), highlighting how population organisation modulates transmission potential.

Whilst the incorporation of clusters we present adds to the literature on epidemic transmission models, there are still simplifications we have made; this study has considered the average cluster size *n* and *x* in the analysis of this model; however, in reality, cluster sizes are different, for example, household sizes vary, even in the same town. Also in this model, the population is assumed to be homogeneous; thus, the socio-demographic heterogeneity that exists in reality has not been included, which may present different sub-populations with different risks of infection. However, natural extensions of epidemic models that incorporate a specific structure that allows between and within-group interactions into epidemic models are available [11]. This novel SeRaMix framework can be extended to these more structured epidemic models, incorporating semi-random mix ideas alongside established and (sometimes) measurable demographic, social, or spatial structures.

The coupling parameter *κ* was fitted to IBM simulations for a single population size (*N* = 1000) and pathogen (*µ* = 0.18, *γ* = 0.16); its generalization across diseases (e.g., airborne vs. contact-based, with varying population sizes) needs further study.

Although this model has not been implemented and tested against real data, the results presented here are aimed at stimulating further research, particularly on the concept of *semi-random mixing* in clustered-interaction epidemic modelling and the public health implications of this model. This framework is hoped to become a useful tool in the hands of epidemiological modellers who should carefully consider this approach and its applicability (while taking note of its limitations) for designing informed, effective public health strategies tailored to reduce or prevent disease transmission within multi-clustered populations.

In the future, this approach could be used to explore heterogeneous cluster sizes in the model simulation and analysis, and better account for socio-demographic heterogeneity, incorporating the differing nature of interactions in and out of households. This model could be extended for more complex dynamics and fitted to specific outbreak data to study the effect of clustered interactions in the design and analysis of NPIs. A detailed investigation of the impact of semi-random interactions could also be done using other modelling methods.

## Conclusion

This study presents a novel theoretical framework for epidemic modelling that introduces the concept of *Semi-Random Mixing* in equation-based epidemic modelling to account for transmission dynamics in multi-clustered populations. Building upon classical transmission models, the developed framework incorporates two fundamental aspects of real-world contact patterns: (1) semi-random mixing within localised clusters (e.g., households, classrooms, workplaces), and (2) limited connectivity between clusters (e.g., between household interactions and visitation). This work provides a derivation of a modified transmission term that relaxes the homogeneous mixing assumption of classical compartmental models while maintaining its analytical tractability. The formulation incorporates cluster size (*n*) and external connections (*x*) through a saturation function, capturing how constrained interaction patterns affect disease spread, and we show that this method can lead to altered predictions for the impact of various interventions compared to the classical model formulation.

## Acknowledgments

MLS was funded by the Institute for Global Pandemic Planning (IGPP), Warwick Medical School, University of Warwick, UK.

## Author contributions statement

Conceptualisation: M. L. S. Supervision: A.S. & K. S. R. Software: K. S. R. & M. L. S. Writing: A. S., K. S. R. & M. L. S.

## Data availability

The MATLAB code is available on GitHub https://doi.org/10.5281/zenodo.16864487

## Ethics declarations

This study focuses on the theoretical development of a mathematical model for the spread of a hypothetical infectious disease. No data was collected or analysed.

## Open access

For the purpose of open access, the authors have applied a Creative Commons Attribution (CC-BY) licence to any Author Accepted Manuscript version arising from this submission.

## References

1. Inglesby, T. V., Nuzzo, J. B., O’Toole, T. & Henderson, D. A. Disease mitigation measures in the control of pandemic influenza. Biosecurity Bioterrorism: Biodefense Strateg. Pract. Sci. 4, 366–375 (2006).

2. Fong, M. W. et al. Nonpharmaceutical measures for pandemic influenza in nonhealthcare settings—social distancing measures. Emerg. infectious diseases 26, 976 (2020).

3. Qiu, Z. et al. Understanding the coevolution of mask wearing and epidemics: A network perspective. Proc. Natl. Acad. Sci. 119, e2123355119 (2022).

4. Hadley, L. et al. Challenges on the interaction of models and policy for pandemic control. Epidemics 37, 100499 (2021).

5. Moein, S. et al. Inefficiency of SIR models in forecasting covid-19 epidemic: a case study of isfahan. Sci. reports11, 4725 (2021).

6. Eker, S. Validity and usefulness of COVID-19 models. Humanit. Soc. Sci. Commun. 7 (2020).

7. Ioannidis, J. P., Cripps, S. & Tanner, M. A. Forecasting for covid-19 has failed. Int. journal forecasting 38, 423–438 (2022).

8. Kretzschmar, M. E. et al. Challenges for modelling interventions for future pandemics. Epidemics 38, 100546 (2022).

9. Johnson, S. Epidemic modelling requires knowledge of the social network. J. Physics: Complex. 5, 01LT01 (2024).

10. Keeling, M. J. & Eames, K. T. Networks and epidemic models. J. royal society interface 2, 295–307 (2005).

11. Inaba, H. Age-structured population dynamics in demography and epidemiology, vol. 1 (Springer, 2017).

12. Science Media Centre. Expert reaction to a study claiming that epidemic modelling needs to be overhauled to include social networks. https://www.sciencemediacentre.org/expert-reaction-to-a-study-claiming-that-epidemic-modelling-needs-to-be-overhauled-to-include-social-networks/ (2024). Accessed: 2024-12-31.

13. Keeling, M. The implications of network structure for epidemic dynamics. Theor. population biology 67, 1–8 (2005).

14. Begon, M. et al. A clarification of transmission terms in host-microparasite models: numbers, densities and areas. Epidemiol. & Infect. 129, 147–153 (2002).

15. McCallum, H., Barlow, N. & Hone, J. How should pathogen transmission be modelled? Trends ecology & evolution 16, 295–300 (2001).

16. Grassly, N. C. & Fraser, C. Mathematical models of infectious disease transmission. Nat. Rev. Microbiol. 6, 477–487 (2008).

17. De Jong, M. C., Diekmann, O. & Heesterbeek, H. How does transmission of infection depend on population size. Epidemic models: their structure relation to data 84 (1995).

18. Mollison, D. Epidemic models: their structure and relation to data. 5 (Cambridge University Press, 1995).

19. Thrall, P. H., Biere, A. & Uyenoyama, M. K. Frequency-dependent disease transmission and the dynamics of the silene-ustilago host-pathogen system. The Am. Nat. 145, 43–62 (1995).

20. Halloran, M. E., Longini, I. M., Struchiner, C. J. & Longini, I. M. Design and analysis of vaccine studies, vol. 18 (Springer, 2010).

21. Keeling, M. J. & Rohani, P. Modeling Infectious Diseases in Humans and Animals (Princeton University Press, 2011).

22. Rock, K., Brand, S., Moir, J. & Keeling, M. J. Dynamics of infectious diseases. Reports on Prog. Phys. 77, 026602 (2014).

23. An, L. et al. Challenges, tasks, and opportunities in modeling agent-based complex systems. Ecol. Model. 457, 109685 (2021).

24. Bonabeau, E. Agent-based modeling: Methods and techniques for simulating human systems. Proc. national academy sciences 99, 7280–7287 (2002).

25. Li, J., Li, C. & Li, X. Quantifying the contact memory in temporal human interactions. In 2018 IEEE International Symposium on Circuits and Systems (ISCAS), 1–5 (IEEE, 2018).

26. Miritello, G., Lara, R., Cebrian, M. & Moro, E. Limited communication capacity unveils strategies for human interaction. Sci. reports 3, 1950 (2013).

27. Ubaldi, E. et al. Asymptotic theory of time-varying social networks with heterogeneous activity and tie allocation. Sci. reports 6, 35724 (2016).

28. Chen, W. Explosive percolation in random networks (Springer, 2014).

29. Grimmett, G. & Grimmett, G. What is percolation? (Springer, 1999).

30. Erdos, P., Rényi, A. et al. On the evolution of random graphs. Publ. math. inst. hung. acad. sci 5, 17–60 (1960).

31. Bollobás, B. The Evolution of Random Graphs—the Giant Component, 130–159. Cambridge Studies in Advanced Mathematics (Cambridge University Press, 2001).

32. Newman, M. Random graphs. In Networks, DOI: 10.1093/oso/9780198805090.003.0011 (Oxford University Press, 2018). https://0-academic-oup-com.pugwash.lib.warwick.ac.uk/book/0/chapter/203818048/chapter-pdf/43641069/oso-9780198805090-chapter-11.pdf.

33. Chowell, G. Fitting dynamic models to epidemic outbreaks with quantified uncertainty: A primer for parameter uncertainty, identifiability, and forecasts. Infect. Dis. Model. 2, 379–398 (2017).

34. Earn, D. J., Park, S. W. & Bolker, B. M. Fitting epidemic models to data: A tutorial in memory of fred brauer. Bull. Math. Biol. 86, 109 (2024).

35. The MathWorks, Inc. Optimization Toolbox User’s Guide. The MathWorks, Inc., Natick, MA, version 2 edn. (2001). fminsearch function.

36. Diekmann, O., Othmer, H. G., Planqué, R. & Bootsma, M. C. On discrete time epidemic models in kermack-mckendrick form. MedRxiv 2021–03 (2021).

37. Isham, V. Stochastic models for epidemics. Oxf. statistical science series 33, 27 (2005).

38. Yaga, S. J. & Saporu, F. W. A study of a stochastic model and extinction phenomenon of meningitis epidemic. Epidemiol. Methods 14, 20240015 (2025).

39. Lloyd, A. L., Zhang, J. & Root, A. M. Stochasticity and heterogeneity in host–vector models. J. Royal Soc. Interface 4, 851–863 (2007).

40. Gov.UK. Making a support bubble with another household. https://www.gov.uk/guidance/making-a-support-bubble-with-another-household (2023). Accessed: 9-November-2024.

41. Levins, R. & Culver, D. Regional coexistence of species and competition between rare species. Proc. Natl. Acad. Sci. 68, 1246–1248 (1971).

42. Hanski, I. Metapopulation dynamics. Nature 396, 41–49 (1998).

43. Cota, W., Soriano-Paños, D., Arenas, A., Ferreira, S. C. & Gómez-Gardeñes, J. Infectious disease dynamics in metapopulations with heterogeneous transmission and recurrent mobility. New J. Phys. 23, 073019 (2021).

