## Supplementary Information for "Modelling Semi-Random Human Mixing: A Modified Force of Infection in Population-level Epidemic Modelling"

#### Model Formulation

We consider a population of $N$ individuals, distributed across $K$ clusters. Each individual in cluster $k$ of size $n_k$ is connected (not to be confused with making contact) to the other $n_k - 1$ members of their cluster. In addition, each person has $x_k$ external connections randomly chosen from the other $K-1$ clusters. For each person, $(n_k - 1) + x_k$ is the number of their connections from which daily contacts are randomly selected. We want to measure how saturated the population is in terms of connectivity of people (accounting for how likely everyone will make contact with everyone else). We consider the average cluster size to be $n$ and the average external connection to be $x$.

##### Critical Threshold for Saturation

To measure the **saturation** of the connections, we use a probabilistic model based on percolation theory [1, 2] and network connectivity [3, 4, 5] to formulate the model. In this model, we consider clusters as **supernodes**, and inter-cluster connections are treated as **edges between supernodes**. This is a similar network structure as the **Erdős–Rényi (ER) random network** $G(K, p)$ [3], where: $K = \frac{N}{n}$ is the number of supernodes (clusters), and $p$ is the probability of an edge (connection) between any two clusters.

Each individual in a cluster forms $x$ external connections randomly to other clusters. Since there are $n$ individuals in each cluster, the expected number of inter-cluster edges (connections) in the population is:

$$\text{Expected inter-cluster edges (connections)} = \frac{n \cdot x \cdot K}{2} = \frac{Nx}{2}. \tag{1}$$

The division by 2 is due to the un-directedness of the connections between people as we do not want to count each connection between a pair of people twice; if a pair of people have a contact which can result in the infection of one of them it is likely that the type of contact could lead to an infection in either direction, depending on the current infection state of each person. In an ER graph, the number of edges is $\binom{K}{2} p \approx \frac{K^2 p}{2}$. Equating the two:

$$\frac{Nx}{2} \approx \frac{K^2 p}{2} \implies p \approx \frac{Nx}{K^2} = \frac{xn}{K} = \frac{xn^2}{N}. \tag{2}$$

In the supernode ER graph, the average degree is:

$$\langle d \rangle = p(K-1) \approx \frac{xn^2}{N} \cdot \frac{N}{n} = xn. \tag{3}$$

assuming that $K \gg 1$ so the $K - 1 \approx K$ or $\frac{N}{n} - 1 \approx \frac{N}{n}$.

To quantify the level of connectivity to allow pathogen spread through the population, we will utilise two known characteristics of ER random graphs: the giant component and the full connectivity threshold. From the connectivity threshold in the Erdős–Rényi (ER) model [3, 4], for a graph (population) with $K$ nodes (clusters), the network becomes connected (no isolated supernodes or clusters) with high probability when the number of edges $E$ satisfies:

$$E \geq \frac{K \ln K}{2}. \tag{4}$$

Since Total inter-cluster links (connections) $(E) = \frac{nx \cdot K}{2} = \frac{Nx}{2}$, the critical condition for connectivity is:

$$\frac{Nx}{2} \geq \frac{K \ln K}{2}. \tag{5}$$

This is written as:

$$\frac{K \cdot n \cdot x}{2} \geq \frac{K \ln K}{2}. \tag{6}$$

Solving for the average supernode degree, $nx$ gives:

$$nx = \langle d \rangle \geq \ln(K) = \ln\left(\frac{N}{n}\right). \tag{7}$$

Thus, the **critical cluster connections** required for the population to be connected is:

$$\tilde{C} = \ln(K) \tag{8}$$

Here, for a fixed population size $N$, the right-hand side, $\ln\left(\frac{N}{n}\right)$, depends on the average cluster size $n$. As $n$ increases, the threshold for connectivity becomes easier to satisfy because $\ln\left(\frac{N}{n}\right)$ decreases. This makes sense because more people are already connected within the clusters, and since there are fewer clusters as $K = \frac{N}{n}$ is reduced, inter-cluster connectivity is easily achieved. Thus, two separate populations of the same size, but with different average cluster sizes, will have different levels of connectivity, which will impact the speed of spread of infectious diseases. For fixed $n$ and $x$, $\ln(K)$ increases as $K$ increases. In this case, $K$ increases with a larger population size, $N$. This will make the threshold for population connectivity (equation (7)) less likely to be satisfied as compared to a smaller population. For a fixed cluster size, $n$, and external contact size $x$, as we increase $K$ by adding more clusters (increasing $N$), it becomes less likely that the critical cluster connection threshold is met. Thus, if we assume $nx$ is fixed, the right-hand side, $\ln(K)$ which does not explicitly depend on $n$, increases as $K$ increases, making the threshold for connectivity difficult or slower to satisfy because $\ln(K)$ increases. This means that two populations with the same average connectivity, $nx$ will have different likelihood of everyone knowing everyone else, depending on the total population. This makes sense because people in a population with fewer clusters are likely to know each other and can spread pathogens faster than in a different population with more clusters if they both have the same connectivity, $nx$.

In the classical Erdős–Rényi model $G(K,p)$, each of the $\binom{K}{2}$ possible edges is included independently with probability $p$. Due to this independent-edge construction, *"the degree of an individual vertex or node of $G(K,p)$ is a Binomial random variable with parameters $K-1$ and $p$ [6]"*. The Binomial characterization yields the mean of the degree distribution,

$$\langle d \rangle \;=\; p(K-1), \tag{9}$$

and also characterizes the model's threshold phenomena—most notably the emergence of a giant component when $p(K-1) \approx 1$ and the graph's near-sure connectivity once $p(K-1) \gtrsim \ln(K)$ [3, 4]. In the theory of random graphs, the *giant component* is the unique largest connected subgraph whose size grows linearly with the number of vertices (clusters in our model).

Assuming that $\theta$ is the fraction of clusters in the population that do not belong to the giant component, for a specific cluster $k$ not to belong to the giant component, the cluster $k$ should not connect to another cluster $k^* \in K-1$ that is connected to the giant component, or if cluster $k$ is connected to $k^*$, then $k^*$ itself should not be connected to the giant component. For external connectivity, each cluster (say cluster $k$) has $K-1$ other clusters to choose from. Since cluster $k$ has $nx$ possible attempts at connecting to any other cluster, the fraction $\frac{nx}{K-1}$ is the probability that cluster $k$ will connect to any particular cluster in the population via its $nx$ external connections. Hence, $1-\frac{nx}{K-1}$ is the probability that none of the $nx$ external connections is connected to cluster $k^*$. Following a similar method in the literature [5], the probability that $k$ is connected to $k^*$, which itself is not a member of the giant component, is $\frac{nx}{K-1}\theta$. Thus, the total probability that $k$ is not connected to the giant component through cluster $k^*$ is $1-\frac{nx}{K-1}+\frac{nx}{K-1}\theta$. Since there are $K-1$ possible clusters through which $k$ can get connected to the giant component, the probability that a cluster $k$ is not connected to the giant component is $\left(1-\frac{nx}{K-1}+\frac{nx}{K-1}\theta\right)^{K-1}$. But recall that this probability is $\theta$. This gives the following implicit relationship:

$$\theta = \left(1-\frac{nx}{K-1}(1-\theta)\right)^{K-1}. \tag{10}$$

Considering $K$ to be large relative to $nx$ so that $\frac{nx}{K-1}$ is small. Thus, using the approximation $1-z \approx e^{-z}$ for small $z$, equation (10) can be rewritten as:

$$\theta \;=\; \lim_{K\to\infty}\left(1-\frac{nx}{K-1}(1-\theta)\right)^{K-1} = e^{-nx(1-\theta)}. \tag{11}$$

This is the self-consistent expression for $\theta$ similar to the ones in the literature [5].

By taking the complementary probability— $\Gamma = 1-\theta$—the probability that a cluster $k$ is connected to the giant component is given as:

$$\Gamma \;=\; 1-e^{-nx\Gamma}. \tag{12}$$

This is the fraction of nodes (clusters) in the largest connected subgraph of a random graph with $K$ nodes (clusters) with average degree (number of connections per cluster) $nx$, [3, 5].

Since the goal of this work is not only to be able to quantify the size of the giant component, but also to approximate the connectedness of the population. We consider a modification to equation (12).

The proportion of the population in the giant component $\Gamma$ reflects the subset of individuals ($\Gamma \times N$) that act as drivers of inter-connectedness in the population. Here, equation (12) contains $\Gamma$ in its exponent; as such, its solution is complex. Without loss of generality, we introduce an approximation to the $\Gamma$, so that henceforth, the notation of the approximated function changes, and is written as:

$$\mathscr{S} \;=\; 1-e^{-nx}. \tag{13}$$

The inequality $1-e^{-nx} \geq 1-e^{-nx\Gamma}$ holds, since $0 \leq \Gamma \leq 1$. Hence, equation (13) does not hold the same meaning as the size (in proportion) of the giant component in its strict sense. We will utilise $\mathscr{S}$ to define a "saturation" function, which will be used directly in our model to capture the connectivity of the network.

The exponential form $1-e^{-y}$ (as in equation (13)) appears in percolation theory [7, 8, 4] to model the probability of global connectivity, as it naturally interpolates between disconnected ($y \to 0$) and connected ($y \to \infty$) phases.

The approximated function (equation (13)) can be written as $\mathscr{S} \;=\; 1-e^{-\langle d\rangle}$. We will analyse it against two critical network properties: *full connectivity*, achieved when $\langle d\rangle \geq \ln(K)$ (ensuring all clusters are interlinked), and the *giant component*, which emerges at $\langle d\rangle \geq 1$ (guaranteeing a connected subgraph spanning a finite fraction of the population). From equation (8), the critical value of $x$ required to achieve full connectivity is $x = \frac{1}{n}ln(N/n)$. From this relationship, as $n$ increases, the value of $x$ required to have full connectivity within the cluster decreases, and vice versa, because an increase in $n$ ensures greater connectivity within the clusters. However, the critical value of $n$ required to achieve full connectivity is $n = \frac{1}{x}ln(N/n)$. This is a self-consistent equation for $n$, because $n$ appears on both sides, where its solution is analytically complex because it requires finding an $n$ that satisfies the equation with itself involved.

From the conditions for full connectivity in Equation (7), the average degree of cluster required is given by substituting for $n$ and $x$ as:

$$\langle d\rangle^* \;=\; \frac{\left(ln(N/n)\right)^2}{nx}. \tag{14}$$

Equation (14) is the minimum average degree required for full cluster connectivity. Thus, for $\langle d\rangle \geq \langle d\rangle^*$, contacts in the population are saturated with little or no differences in $\mathscr{S}$ regardless of how much $\langle d\rangle$ increases. To ensure saturation behavior, $\langle d \rangle = nx$ in equation (13) is weighted by a function $\frac{\kappa}{n+1}$, where, $\kappa$ is a coupling constant that acts like a scaling factor or effectiveness coefficient relating how strongly the cluster dynamics (internal/external) are influenced by changing the population configuration $(n, x, N)$. The values of $\kappa$ can be estimated from empirical data. This leads to equation (15), which will henceforth be referred to in this work as the *'saturation function'*.

$$\mathscr{S} = 1 - e^{-\kappa \frac{xn}{n+1}}. \tag{15}$$

The reasons for choosing $\frac{\kappa}{n+1}$ are: it has the lowest degree polynomial ratio (rational function), the fewest parameters, it is monotonic increasing, it facilitates saturation when multiplied by $nx$, and it does not have any additional complexity like powers, exponentiations, or higher degree polynomials.

Since the introduction of $\frac{\kappa}{n+1}$ is aimed at reducing $\mathscr{S}$, the upper value of $\kappa$ is 1. This will ensure that the ratio satisfies $0 \leq \frac{\kappa}{n+1} \leq 1 \; \forall \, n \geq 1$.

Using an individual-based model (IBM) to simulate the spread of an epidemic in clustered populations will help validate this formulation and estimate $\kappa$. In this model, each person is given their potential contacts, which are made up of both intra-cluster connections (controlled by cluster size $n$) and inter-cluster connections (controlled by $x$). Each person chooses their daily contacts randomly from their assigned neighbours. Good agreement between the predicted model with $\mathscr{S}$ and the IBM simulated outbreak dynamics for different $n$ and $x$ values will confirm that the saturation function $\mathscr{S}$ correctly quantifies how likely that everyone will meet with everyone else in populations with different average cluster sizes, $n$, and the average external connections, $x$.

From equation (15), when $\kappa xn \ll n+1$, $\mathscr{S} \approx 0$, corresponds to fragmented clusters with no giant component. When $\kappa xn \gg n+1$, $\mathscr{S} \approx 1$, matching the full connectivity regime. The transition between these extremes mirrors real-world connectivity that we aim to quantify as suitable for different average cluster sizes, $n$, and the average per-capita external connections, $x$. This ratio quantifies how "far" the population is from achieving contact saturation.

#### Probability of disease transmission (Non-linear Force of Infection)

For the rest of this work (except where indicated), we will present the per susceptible force of infection explicitly in terms of the key quantities described earlier rather than the traditional approach of writing it as $\beta \frac{I(t)}{N}$. Thus, we will reformulate the force of infection in terms of $\mu$ (which is pathogen-dependent), the contact rate $c$, and the per-contact probability of choosing an infected person, which is given as:

$$p(t) = \frac{I(t)}{N}\left(1 - e^{-\kappa \frac{xn}{n+1}}\right). \tag{16}$$

This approach would enable me to distinguish the behavioural/social constituents ($c$, $n$, and $x$) in the force of infection from the biological quantity, $\mu$, in the analysis of non-pharmaceutical intervention.

In this formulation, the per-contact probability that a susceptible person will be infected at time $t$ is given by $\mu p(t)$, for $c = 1$. However, a person may have more than one contact per day, as such to accommodate this, first take the complementary probability to write the probability that a contact will not lead to infection as $1 - \mu p(t)$, given that $\mu p(t)$ is the probability of infection per contact. If a susceptible person contacts $c$ people of unknown infectious status at time $t$, then the probability of not being infected after these contacts is:

$$Q(t) = \Big(1 - \mu p(t)\Big)^c. \tag{17}$$

This means that the probability of no transmission would have occurred $c$ times at time $t$ [9]. Thus, applying another complementary probability, the probability that a susceptible person will be infected after contact with $c$ persons of unknown infectious statuses at any time $t$ is given as:

$$P(t) = 1 - Q(t) = 1 - \Big(1 - \mu p(t)\Big)^c. \tag{18}$$

Thus, $P(t)$ is the force of infection equivalent to $\beta p(t)$ that drives the spread of diseases.

#### Algorithms

For IBM, we generated $N = 1,000$ individuals and randomly assigned each to a cluster to mimic random local interactions at work or home. For each individual, we assign $x$ randomly selected individuals from other clusters. For each individual, we assigned their $n-1$ cluster neighbours and the $x$ randomly selected individuals as their social circle (these are the potentially recurrent contacts of each individual) so that each person has $n-1+x$ connections out of which they randomly select their $c$ daily contacts at each time step. We used the $SIR$-type IBM, analogous to the SeRaMix-EBM presented in the main text, which is outlined in Algorithm 1.

**Algorithm 1** Run SIR Semi-Random Mixing IBM

**Input:** Total population $N$, per-contact transmission probability $\mu$, recovery probability $\gamma$, number of daily contacts $c$, average cluster size $n$, average external connections $x$.
**Output:** Counts of susceptible ($S$), infected ($I$), and removed ($R$) individuals at each time step

**Initialization:**
Set all $N$ individuals as susceptible ($S \leftarrow N$, $I \leftarrow 0$, $R \leftarrow 0$)
Randomly select one individual and set their status to infected ($I \leftarrow 1$, $S \leftarrow S-1$)
Assign each individual $n-1$ internal and $x$ external connections. This forms the potentially recurrent contacts for each individual
Initialize arrays to store $S$, $I$, and $R$ counts over time

**for** each time step $t = 0, 1, \ldots$ MaxTime **do**
  **for** each individual $i$ in the population **do**
    **Contact Selection:**
    Randomly select $c$ contacts from $i$'s $n-1+x$ potentially recurrent contacts without replacement
    Count the number of infected contacts (*InfectedContacts*) out of the $c$ contacts

    **Infection Dynamics:**
    **if** individual $i$ is susceptible **then**
      Compute force of infection: $foi \leftarrow \mu \cdot InfectedContacts$
      Generate random number $r \in [0,1]$
      **if** $r < foi$ **then**
        Set individual $i$ as infected
        Update counts: $S \leftarrow S-1$, $I \leftarrow I+1$
      **end if**
    **end if**

    **Recovery Dynamics:**
    **if** individual $i$ is infected **then**
      Generate random number $r \in [0,1]$
      **if** $r < \gamma$ **then**
        Set individual $i$ as removed (recovered)
        Update counts: $I \leftarrow I-1$, $R \leftarrow R+1$
      **end if**
    **end if**
  **end for**

  **Record State:**
  Store counts of $S$, $I$, and $R$ for time $t$
**end for**

**Algorithm 2** Parameter Estimation for Discrete SIR-Type Semi-Random Mixing EBM

**Input:**
  Incidence datasets $Inc_{data}(n,x,r)$ for $n \in \{4,5,10,20\}, x \in \{1,\ldots,6\}, r \in \{1,\ldots,R\}$,
  $N$ (total population), $\mu$ (transmission probability),
  $C$ (average contacts), $\gamma$ (recovery probability),
  $R$ (number of realizations)
**Output:**
  $\kappa$ (single estimate for all datasets)
  Model fits for plotting
**Initialization:**
  Load all datasets $Inc_{data,all}(n,x,r)$
  Initialize time vector $t_{\text{data}}$
  Set optimization bounds: $\kappa \in [0,\infty)$
  Set initial guess $\kappa^{(0)}$ (scalar)
**Data Preparation:**
Initialize empty lists $Inc_{\text{data, all}}$, $x_{\text{all}}$, $n_{\text{all}}$
**for** each cluster size $n \in \{4,5,10,20\}$ **do**
  **for** each external connection $x = 1$ to 6 **do**
    **for** each realization $r = 1$ to $R$ **do**
      $Inc_{\text{data}} \leftarrow Inc_{data,all}(n,x,r)$ ▷ Extract incidence
      **if** $Inc_{\text{data}}$ is not all zeros **then**
        Append $Inc_{\text{data}}$ to $Inc_{\text{data, all}}$
        Append $x$ to $x_{\text{all}}$, $n$ to $n_{\text{all}}$
      **else**
        Skip realisation
      **end if**
    **end for**
  **end for**
**end for**
$\kappa \leftarrow \text{lsqnonlin}(\text{residual\_fn}, \kappa^{(0)}, \text{lb}, \text{ub}, Inc_{\text{data, all}}, x_{\text{all}}, n_{\text{all}}, N, \mu, C, \gamma)$ ▷ Fit single $\kappa$
**Model Fits:**
**for** each dataset $(n,x)$ **do**
  Simulate SIR model with $\kappa$, $n$, $x$, $N$, $\mu$, $C$, $\gamma$
  Compute model incidence $Inc_{\text{model}}$
  Store $Inc_{\text{model}}$ for plotting against $Inc_{\text{data, mean}}(n,x)$
**end for**
**Return:** $\kappa$, model fits

**Algorithm 3** Residual Function for Discrete SIR-Type Semi-Random Mixing EBM

**function** RESIDUAL_FN($\kappa$, $Inc_{\text{data, all}}$, $x_{\text{all}}$, $n_{\text{all}}$, $N$, $\mu$, $C$, $\gamma$)
  **Input:**
    $\kappa$ (single scalar parameter)
    $Inc_{\text{data, all}}$ (list of all incidence data)
    $x_{\text{all}}$ (external connections for each data)
    $n_{\text{all}}$ (cluster sizes for each data)
    $N$ (total population), $\mu$ (transmission probability),
    $C$ (average contacts), $\gamma$ (recovery probability)
  Initialize empty all_residuals
  **for** each dataset $idx = 1$ to length($Inc_{\text{data, all}}$) **do**
    Get $x \leftarrow x_{\text{all}}(idx)$, $n \leftarrow n_{\text{all}}(idx)$
    $Inc_{\text{data}} \leftarrow Inc_{\text{data, all}}\{idx\}$ ▷ Get incidence data
    num_points ← length($Inc_{\text{data}}$)
    $I_{\text{model}}(1:\text{num\_points}) \leftarrow 0$, $S_{\text{model}}(1:\text{num\_points}) \leftarrow 0$
    $I_{\text{model}}(1) \leftarrow 1$, $S_{\text{model}}(1) \leftarrow N - I_{\text{model}}(1)$
    $P_t(1:\text{num\_points}-1) \leftarrow 0$ ▷ Initialize infection probability
    **for** $t = 1$ to num_points − 1 **do**
      factor $\leftarrow 1 - e^{-\kappa \cdot x \cdot n/(n+1)}$ ▷ Compute saturation function
      $P_t(t) \leftarrow 1 - \left(1 - \mu \cdot \frac{I_{\text{model}}(t)}{N} \cdot \text{factor}\right)^C$ ▷ Compute infection probability
      $S_{\text{model}}(t+1) \leftarrow S_{\text{model}}(t) - S_{\text{model}}(t) \cdot P_t(t)$ ▷ Update susceptible
      $I_{\text{model}}(t+1) \leftarrow I_{\text{model}}(t) + S_{\text{model}}(t) \cdot P_t(t) - \gamma \cdot I_{\text{model}}(t)$ ▷ Update infected
    **end for**
    $Inc_{\text{model}} \leftarrow S_{\text{model}}(1:\text{num\_points}-1) \cdot P_t$ ▷ Compute model incidence
    all_residuals ← [all_residuals, $Inc_{\text{data}} - Inc_{\text{model}}$] ▷ Append residuals
  **end for**
  **Return:** all_residuals
**end function**

#### The Reproduction Number

To derive the basic reproduction number, we will begin by linearising the force of infection $P(t)$. We used binomial expansion, which states that for any real number $f$ and positive integer $m$, the following holds:

$$(1+f)^m = 1 + mf + (m(m-1)f^2)/2! + (m(m-1)(m-2)f^3)/3! + \ldots \tag{19}$$

Noting that at the beginning of outbreaks, we have $t \to 0$, and the per-contact probability of choosing an infected person, $p(0) << 1$, since we start with a very small fraction of the population initially infected. Following the binomial expansion in equation (19), we may therefore ignore the second and higher orders, and using a similar approach explored in other literature [10, 11, 12], the linearised form of equation (18) is given as:

$$P(t) = 1 - \left(1 - \mu p(t)\right)^c = 1 - \left(1 - \mu c p(t) + \mathscr{O}\left(p(t)^2\right)\right) \approx \mu c p(t). \tag{20}$$

The reproduction number is computed using the next-generation matrix formula for the discrete-time model [13, 14],

$$\mathscr{R}_t = \rho\left(F[\mathscr{I} - T]^{-1}\right). \tag{21}$$

Where $\mathscr{I}$ is the identity matrix, $F$ is the matrix associated with the creation of new infections (transmissions), and $T$ is the matrix associated with the transition from the infected compartments. The term $\rho\left(F[\mathscr{I} - T]^{-1}\right)$ is the spectral radius (defined as the dominant eigenvalue) of the matrix $F[\mathscr{I} - T]^{-1}$, where the matrix of transitions satisfies $\rho(T) < 1$[14].

For the $SIR$-type model (**??**), the infected compartment $I$ is used to derive the reproduction number. Thus, the transmission and transition terms in the compartments are given as:

$$\mathscr{F} = \quad S(t)P(t) = \quad S(t)\mu c \frac{I(t)}{N}\left(1 - e^{-\kappa \frac{xn}{n+1}}\right). \tag{22}$$

$$\mathscr{T} = (1-\gamma)I(t). \tag{23}$$

The Jacobian matrices of transmission and transition are given respectively by partial derivatives of $\mathscr{F}$ and $\mathscr{T}$ with respect to $I$ as:

$$F = S(t)\frac{\mu c}{N}\left(1 - e^{-\kappa \frac{xn}{n+1}}\right). \tag{24}$$

$$T = 1 - \gamma. \tag{25}$$

The condition $\rho\left(T\right) < 1$ is satisfied since $0 < \gamma < 1$. Furthermore,

$$\mathscr{I} - T = \gamma. \tag{26}$$

$$\left[\mathscr{I} - T\right]^{-1} = \frac{1}{\gamma} \tag{27}$$

The next-generation matrix is computed as:

$$F\left[\mathscr{I} - T\right]^{-1} = S(t)\frac{\mu c}{N}\left(1 - e^{-\kappa \frac{xn}{n+1}}\right) \times \frac{1}{\gamma}. \tag{28}$$

The reproduction number is thus given by the maximum absolute value of its eigenvalues as:

$$\rho\left(F[\mathscr{I}-T]^{-1}\right) = \max\left\|S(t)\frac{\mu c}{\gamma N}\left(1-e^{-\kappa\frac{xn}{n+1}}\right)\right\|. \tag{29}$$

Since there is only one term in the next generation matrix, the time-dependent (effective) reproduction number is given by

$$\mathscr{R}_t = S(t)\frac{\mu c}{\gamma N}\left(1-e^{-\kappa\frac{xn}{n+1}}\right). \tag{30}$$

At the beginning of outbreaks $t \to 0$, the number of infected individuals are considered small, and $S(0) \approx N$ so that the basic reproduction number is written as:

$$\mathscr{R}_0 = \frac{\mu c}{\gamma}\left(1-e^{-\kappa\frac{xn}{n+1}}\right) = \frac{\beta}{\gamma}\left(1-e^{-\kappa\frac{xn}{n+1}}\right). \tag{31}$$

Similar to other epidemic models, the basic reproduction number in this model depends primarily on two epidemiological quantities: the infectious rate $\beta$ and the recovery rate $\gamma$. However, this model reveals the additional dependence of $\mathscr{R}_0$ on the saturation function, which is a function of the average cluster size in the population $n$, and the average external connections $x$.

From the knowledge that if the value of $\mathscr{R}_0$ is greater than 1, it signifies that the infection will invade the population, as the average number of secondary infections caused by a single infectious individual in a susceptible population is greater than one [15], the spread of infection in the population is anticipated if the following inequality holds:

$$\mu_T > \frac{\gamma}{c\left(1-e^{-\kappa\frac{xn}{n+1}}\right)}. \tag{32}$$

Equation (32) quantifies the value of biological infectiousness of the pathogen required for an outbreak to occur given that the average cluster size in the population is $n$, the average number of contacts is $c$, the recovery rate is $\gamma$, and the average external connections in the population, $x$. Knowing the threshold quantity is helpful to understand and predict what level of intervention might be required for effective control that requires the failure of the inequality (32). As the biological infectiousness of the pathogen is expected to be constant, reducing the reproduction number below one (failure of this inequality) would require the reduction of all or any of the cluster size $n$ and the number of contacts $c$, the average external connections $x$, or increasing the removal rate $\gamma$ which could be through isolation or hospitalization of those infected.

188 # Additional figures

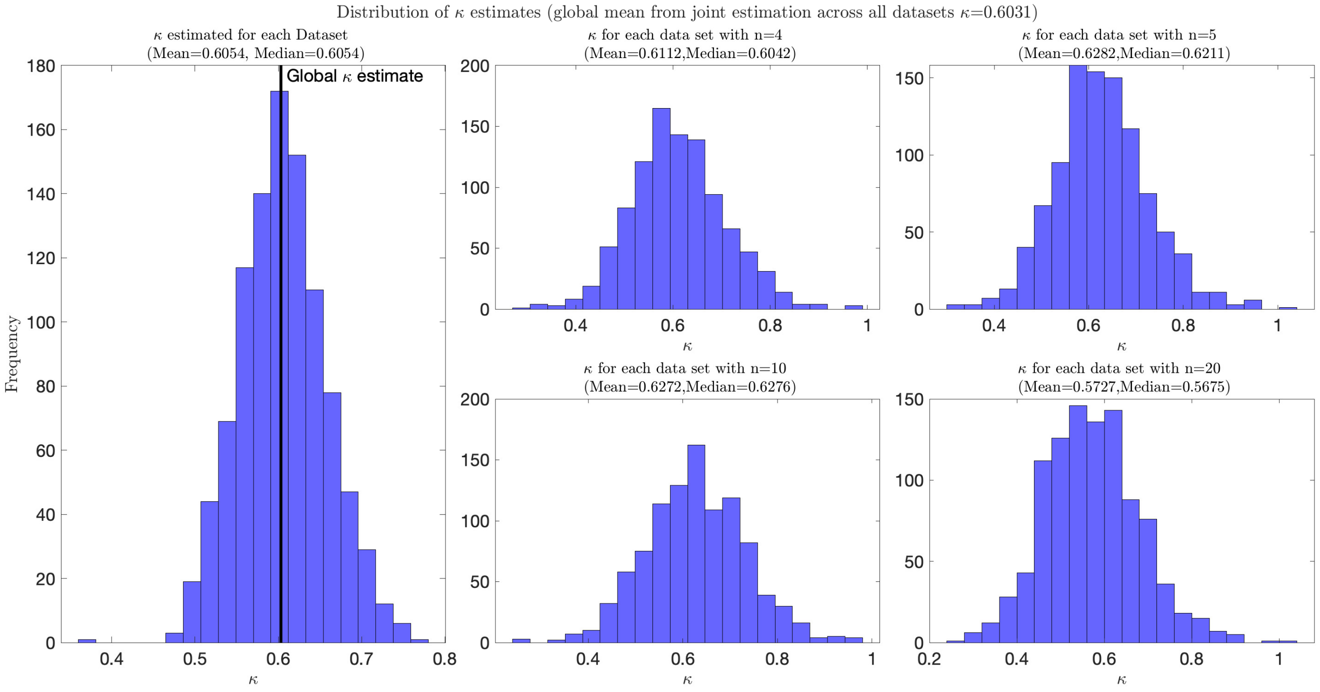


**Figure 1.** Histograms of $\kappa$ estimates from all datasets, and data grouped by cluster sizes $n$. For each $n$ and $x$, 1,000 datasets have been used, yielding 24,000 in total. The left panel shows the distribution of the $\kappa$ fitted jointly across all datasets. The right panels display distributions of $\kappa$ for each cluster size $n = 4, 5, 10, 20$. The overall mean of median $\kappa$ across $n$ and the global mean $\kappa$ for all data jointly fitted are given at the top of the figure.

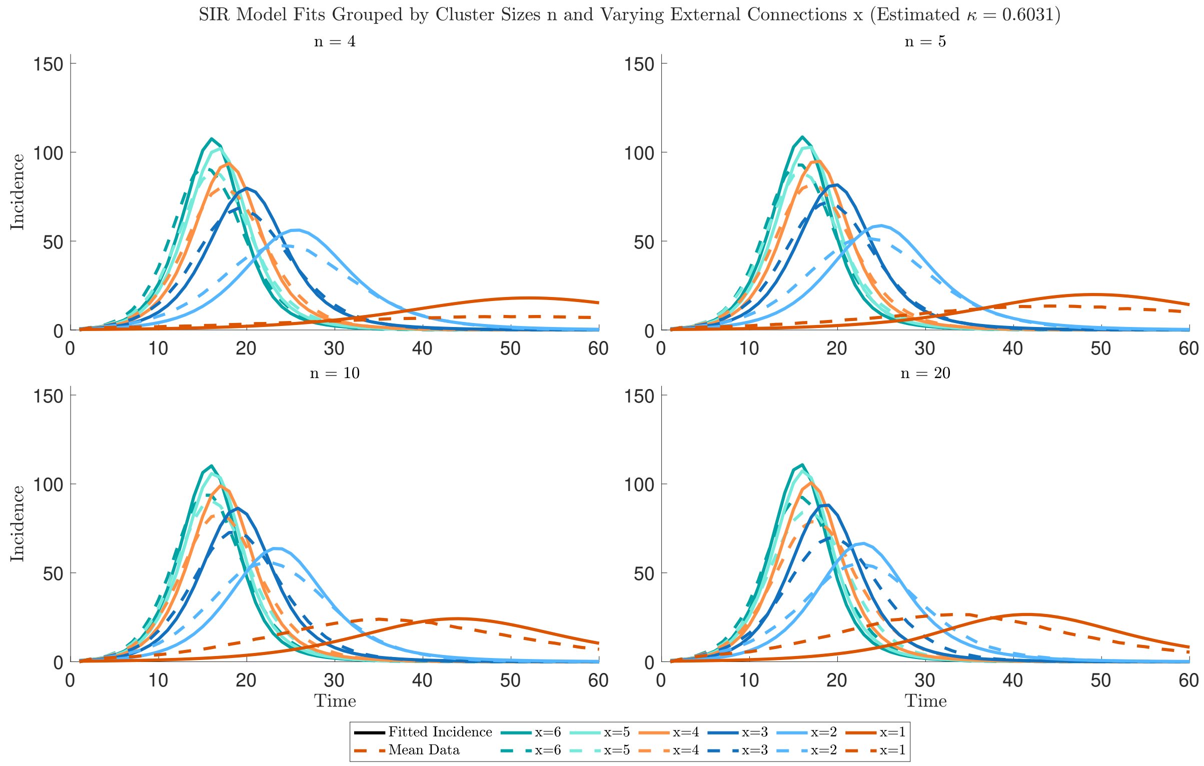


**Figure 2.** EBM fits to the synthetic IBM incidence data (new infections per timestep) grouped by cluster size $n$ with estimated $\kappa$. Each subplot shows infection dynamics for all per-capita external connectivity ($x = 1, \ldots, 6$) at: (a) $n = 4$, (b) $n = 5$, (c) $n = 10$, and (d) $n = 20$. For each $n$ and $x$, 1,000 datasets have been used. Here, I show the mean of the 1,000 IBM simulations used for fitting as a visual representation of the data for comparison purposes. A full summary of the uncertainty from the 1,000 IBM simulations used for the synthetic data is shown in Figure **??**.

### References


1. Chen, W. *Explosive percolation in random networks* (Springer, 2014).
2. Grimmett, G. & Grimmett, G. *What is percolation?* (Springer, 1999).
3. Erdos, P., Rényi, A. *et al.* On the evolution of random graphs. *Publ. math. inst. hung. acad. sci* **5**, 17–60 (1960).
4. Bollobás, B. *The Evolution of Random Graphs—the Giant Component*, 130–159. Cambridge Studies in Advanced Mathematics (Cambridge University Press, 2001).
5. Newman, M. Random graphs. In *Networks*, DOI: 10.1093/oso/9780198805090.003.0011 (Oxford University Press, 2018). https://0-academic-oup-com.pugwash.lib.warwick.ac.uk/book/0/chapter/203818048/chapter-pdf/43641069/oso-9780198805090-chapter-11.pdf.
6. Frieze, A. & Karoński, M. *Introduction to random graphs* (Cambridge University Press, 2015).
7. Stauffer, D. & Aharony, A. *Introduction to percolation theory* (Taylor & Francis, 2018).
8. Meester, R. & Roy, R. *Occupancy in Poisson Boolean models*, 40–90. Cambridge Tracts in Mathematics (Cambridge University Press, 1996).
9. Halloran, M. E., Longini, I. M., Struchiner, C. J. & Longini, I. M. *Design and analysis of vaccine studies*, vol. 18 (Springer, 2010).
10. Soriano-Paños, D., Lotero, L., Arenas, A. & Gómez-Gardeñes, J. Spreading processes in multiplex metapopulations containing different mobility networks. *Phys. Rev. X* **8**, 031039, DOI: 10.1103/PhysRevX.8.031039 (2018).
11. Soriano-Paños, D., Ghoshal, G., Arenas, A. & Gómez-Gardeñes, J. Impact of temporal scales and recurrent mobility patterns on the unfolding of epidemics. *J. Stat. Mech. Theory Exp.* **2020**, 024006, DOI: 10.1088/1742-5468/ab6a04 (2020).
12. Li, J., Lai, S. & Gao, G. F. The emergence, genomic diversity and global spread of sars-cov-2. *Nature* **600**, 408–418 (2021). Https://doi.org/10.1038/s41586-021-04188-6.
13. Allen, L. J. & van den Driessche, P. The basic reproduction number in some discrete-time epidemic models. *J. difference equations applications* **14**, 1127–1147 (2008).
14. Hernandez-Ceron, N., Feng, Z. & Van den Driessche, P. Reproduction numbers for discrete-time epidemic models with arbitrary stage distributions. *J. Differ. Equations Appl.* **19**, 1671–1693 (2013).
15. Diekmann, O., Heesterbeek, J. A. P. & Metz, J. A. On the definition and the computation of the basic reproduction ratio r 0 in models for infectious diseases in heterogeneous populations. *J. mathematical biology* **28**, 365–382 (1990).
